# Efficacy of Tailored Messages for 28-Week Exercise Sustainability in People with HIV

**DOI:** 10.64898/2026.03.29.26349681

**Authors:** Paul F. Cook, Allison R. Webel, Melissa P. Wilson, Christine Horvat Davey, Vitor H. F. Oliveira, Vincent Khuu, Samantha Matzio, Grace L. Kulik, Samantha MaWhinney, Catherine M. Jankowski, Kristine M. Erlandson

## Abstract

People with HIV (PWH) are at increased risk for cardiovascular diseases and other age-related comorbidities. These risks can be reduced through moderate to vigorous physical activity (MVPA), but MVPA can be difficult to sustain over time. We tested tailored text messages combined with motivational interviewing (MI) to sustain MVPA among PWH. Messages were created using Two Minds Theory and matched to daily survey responses about exercise barriers. 118 PWH ages ≥ 50 were initially randomized to high-intensity interval training or continuous moderate-intensity exercise. After 16 weeks, 92 participants were re-randomized to receive either tailored messages plus MI, or educational control messages, for 12 weeks. Both groups completed daily barrier surveys and wore an ActiGraph monitor for 1 week/month. PWH in the tailored-messaging plus MI group maintained their MVPA, ending at *M* = 30.5 minutes per day (*SD* = 36.7), compared to a decrease among PWH in the educational-control group, ending at *M* = 26.0 (*SD* = 25.8), *F*(1, 255) = 5.76, *p* = .02, *d* = 0.51 for the group-by-time interaction using intent-to-treat principles. Findings were similar based on both actigraphy and self-reported MVPA, and were robust to attrition. PWH in the tailored-messaging group also reported higher exercise self-efficacy and better perceived health over time, relative to the educational-control group. Exploratory analyses suggested that the tailored messages’ effects were additive to motivational interviewing. In this study, an automated tailored-messaging intervention led to sustained MVPA. Tailored messages were superior to non-tailored educational messages, and may help PWH maintain their long-term health.

---

People with HIV (PWH) experience accelerated and accentuated aging, meaning that they develop chronic conditions at an earlier age and with greater severity than their peers without HIV [1]. Physical activity can mitigate these risks, especially moderate to vigorous physical activity (MVPA) that is done on a regular basis [2]. Aerobic exercise also has positive effects on mental health for PWH [3]. Despite these benefits of physical activity, many people find it hard to maintain increases in activity over the long term [4], which can weaken the effects of interventions to promote MVPA. Sustaining MVPA may be particularly challenging for PWH, many of whom have lower baseline activity than the general population [5].

## Behavioral Theories for Sustained MVPA

The factors that lead people to initiate a new exercise program can differ from those that promote sustained MVPA over time. Prochaska and colleagues [6] suggested that the most successful initial behavior-change interventions utilize the “strong principle of change,” namely persuasion to increase the perceived importance of the activity. Some examples of strong-principle interventions might include cognitive-behavioral approaches or motivational interviewing (MI). To ensure successful maintenance, however, interventions should focus on the “weak principle of change,” which involves reducing barriers and making the new behavior an unconscious habit [6]. Perhaps because of this distinction, reviews have found inconsistent results for MI in the context of exercise promotion [7]. Kendrick et al. [5] found that social networks are particularly important in maintaining MVPA over time. Similarly, a qualitative study by Maula et al. [8] identified the importance of physical, psychological, social, and environmental factors as barriers or facilitators to sustained MVPA among older adults.

The best available interventions to sustain MVPA among older adults focus on enhancing self-efficacy, self-control, and capability for physical activity [9]. These findings have been interpreted in terms of Bandura’s Social-Cognitive Model, in line with the Behavior Change Consortium’s seminal work on health-behavior maintenance [10]. However, that model’s focus on cognition and attitudes is somewhat inconsistent with Prochaska et al.’s [6] finding that sustaining behavior depends on making it an unconscious habit. A recent expert group [11] instead used Bronfenbrenner’s Socio-Ecological Model, and identified environmental and social factors as more important for sustained MVPA.

A newer theoretical model that emphasizes non-conscious factors – collectively called the “Intuitive Mind” – is Two Minds Theory (TMT: [13]). This model suggests a range of non-conscious influences on behavior including social support, emotions, and sense of identity, in addition to conscious factors – called the “Narrative Mind” – that include attitudes, beliefs, and ideas. TMT posits that the Intuitive Mind controls behavior at any given time, but that the Narrative Mind has a feed-forward capability to affect future Intuitive-level decision-making [14]. For that reason, language-based interventions focused on the conscious Narrative Mind will be most successful in promoting sustained MVPA when their content addresses Intuitive-level factors such as emotions or social perceptions. TMT also suggests that interventions will be more successful when they are delivered in real time and in the context of everyday life, close to when health behaviors occur [13]. TMT is in an integrative model consistent with multiple targets for change and multiple intervention strategies, but differs from most other behavioral theories in predicting that interventions delivered in real time and based on immediate barriers to exercise will have the strongest possible effects on behavior.

## Purpose of the Current Study

We delivered an intervention to sustain MVPA among PWH based on TMT. After the primary intervention phase of a randomized trial comparing high-intensity interval training (HIIT) versus continuous moderate exercise (CME), 92 PWH were re-randomized to receive either tailored text messages or non-tailored educational messages. Tailored text messages were selected to match the participant’s primary barrier to exercise based on a daily electronic survey with 10 possibilities; within each of the 10 barrier categories, there were 25 possible messages, each linked to one of 6 intervention strategies suggested by TMT [13]. TMT-based intervention strategies target factors identified as important in the literature review above, including, e.g., social imagery (a Narrative-mind strategy), problem-solving (also Narrative-level), or use of cues and reminders (an Intuitive-mind strategy). All participants in the tailored-messaging group, and some of those in the educational-control group, also received MI from an exercise coach. The goal of this study was to determine whether tailored messages would help to sustain MVPA among PWH who had already successfully increased their MVPA through a structured exercise program. Our hypothesis was that participants receiving a coaching intervention for self-directed exercise that combines MI and personalized support during the maintenance phase can promote long-term adherence to physical activity. TMT has been used as a theoretical framework to understand patterns of physical activity among PWH [14, 15], but this is the first study to test an intervention specifically designed based on TMT.

## Method

### Participants

Participants were PWH enrolled in the High-Intensity Exercise Study to Attenuate Limitations and Train Habits in Older Adults with HIV (HEALTH: [17]), who completed 16 weeks of supervised exercise. All participants were ≥ 50 years of age, reported fatigue, and had a sedentary lifestyle (MVPA < 3 times per week and no regular resistance exercise) for ≥ 3 months before enrollment. All participants had received HIV antiretroviral treatment (ART) ≥ 1 year, had well-controlled HIV (viral load < 200 copies/mL), owned a cell phone with text-messaging and/or email capabilities, and could speak, read, and write English. Medical exclusion criteria as described in Oliveira et al. [17] were also applied; most relevant to this analysis, participants had no substance use, unstable illness, recent surgery/trauma/injury, or other conditions that in the referring clinician’s judgment would have interfered with participation.

A total of 142 participants were screened between March 9, 2021 and August 28, 2024 from sites in Aurora, CO and Seattle, WA, and 118 participated in structured exercise during phase 1 of the study. Participants came primarily from Ryan White HIV specialty clinics at the University of Colorado and University of Washington. This study was approved by the Colorado Multiple Institutional Review Board (#19-1985) and registered at ClinicalTrials.gov (#NCT04550676), and all participants provided written informed consent. Study flow is shown in Figure 1. A total of 92 phase 1 participants were re-randomized (86 who were included in the phase 1 analysis, plus 6 others who were randomized but exercised independently) to behavioral intervention groups for phase 2 of the study. Participants’ baseline characteristics, overall and by phase 2 group, are presented in Table 1. There were slight between-group differences in gender, education, and employment, but none were statistically significant, suggesting that the re-randomization was effective. Participants started phase 1 with similarly low levels of MVPA, had similar exercise adherence during phase 1 (96.5%), and ended phase 1 with similar levels of MVPA (45.8 minutes/day). Participants also showed similar average levels of improvement in physical functioning [18], frailty indicators [19], and fatigue [20] across phase 1 groups. Phase 2 thus began without any systematic between-group differences; any unexpected sources of variability should have been addressed by re-randomization.

**Fig. 1.**
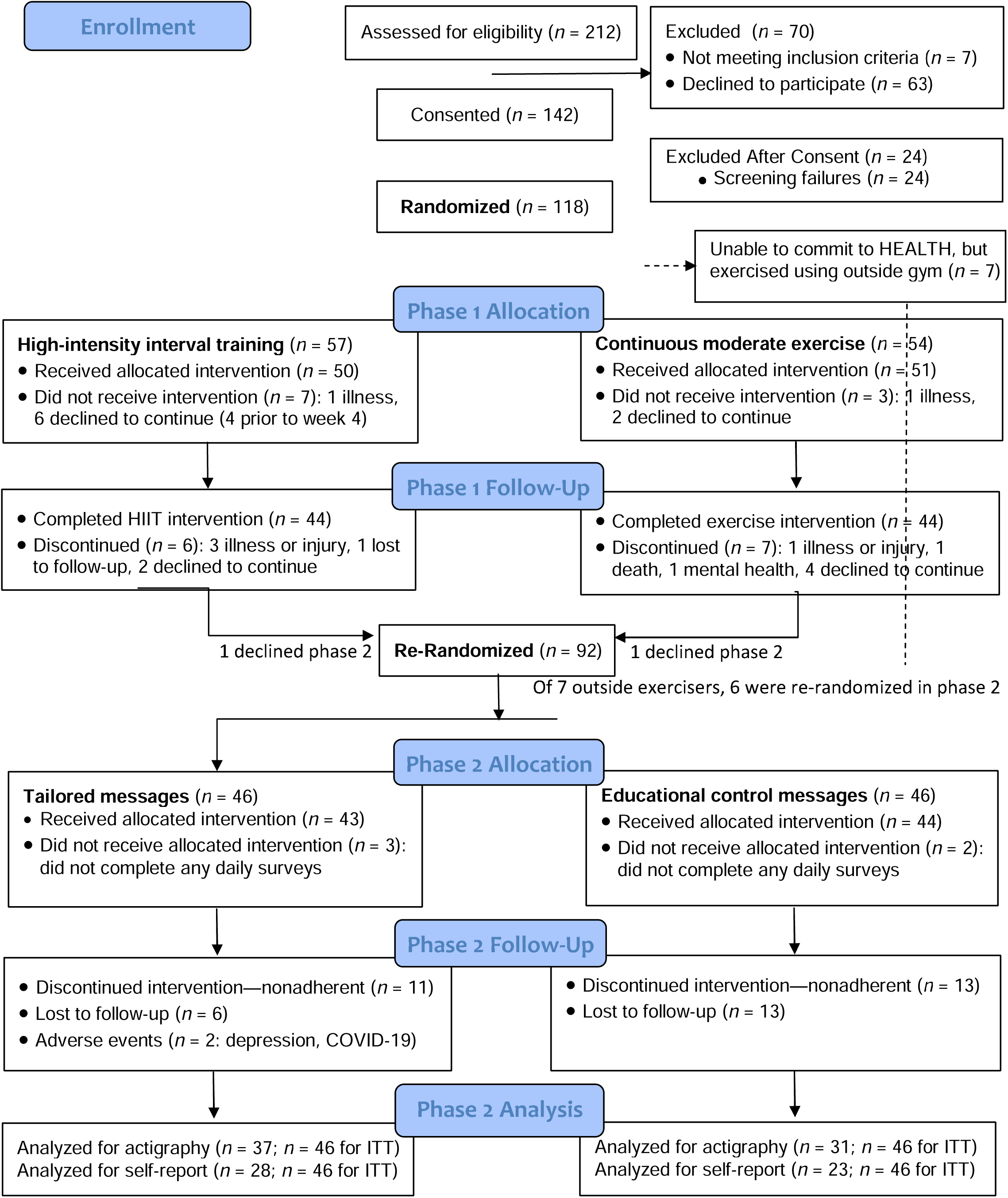
CONSORT Study Flow Diagram

**Table 1.** Participant Demographics.

| Characteristic | N (%) or M (SD) | | | Difference Between Groups<br>(*t* or $\chi^2$, effect size) |
| --- | --- | --- | --- | --- |
|  | Intervention | Control | Full Sample |  |
| Age in Years | 58.5 (0.96) | 59.0 (5.64) | 58.7 (6.04) | $t(90) = -0.36, p = .72, d = 0.08$ |
| Sex at Birth: |  |  |  |  |
| Male | 40 (87.0%) | 39 (84.8%) | 79 (85.9%) | $\chi^2 = 0.90, p = .77, \phi = .10$ |
| Female | 6 (13.0%) | 7 (15.2%) | 13 (14.1%) |  |
| Gender Identity: |  |  |  |  |
| Male | 40 (87.0%) | 35 (76.1%) | 75 (81.5%) | $\chi^2 = 5.67, p = .34, \phi = .25$ |
| Female | 5 (10.9%) | 7 (15.2%) | 12 (13.0%) |  |
| Queer or Nonbinary | 1 ( 2.2%) | 3 ( 6.5%) | 4 ( 4.4%) |  |
| Transgender Male | 0 ( 0.0%) | 1 ( 2.2%) | 1 ( 1.1%) |  |
| Race/Ethnicity: |  |  |  |  |
| Black, non-Hispanic | 7 (15.2%) | 7 (15.2%) | 14 (15.2%) | $\chi^2 = 1.50, p = .68, \phi = .13$ |
| White, non-Hispanic | 31 (67.4%) | 28 (60.9%) | 59 (64.1%) |  |
| Hispanic, any race | 5 (10.9%) | 9 (19.6%) | 14 (15.2%) |  |
| Other or not reported | 3 ( 6.5%) | 2 ( 4.3%) | 5 (11.9%) |  |
| Lowest Reported CD4: |  |  |  |  |
| < 200 cells/mm<sup>3</sup> | 25 (54.3%) | 23 (50.0%) | 48 (52.2%) | $\chi^2 = 4.64, p = .33, \phi = .22$ |
| 200-350 cells/mm<sup>3</sup> | 8 (17.4%) | 6 (13.0%) | 14 (30.4%) |  |
| 351-500 cells/mm<sup>3</sup> | 4 ( 8.7%) | 2 ( 4.3%) | 6 ( 6.5%) |  |
| > 500 cells/mm<sup>3</sup> | 2 ( 4.3%) | 8 (17.4%) | 10 (10.9%) |  |
| Not sure | 7 (15.2%) | 7 (15.2%) | 14 (15.2%) |  |
| VACS Score | 33.2 (9.66) | 33.5 (9.48) | 33.3 (9.52) | $t(89) = 0.14, p = .89, d = 0.03$ |
| BMI Score | 30.0 (6.72) | 30.2 (5.70) | 30.1 (6.20) | $t(90) = 0.19, p = .85, d = 0.04$ |
| Level of Education: |  |  |  |  |
| 11<sup>th</sup> grade or less | 1 ( 2.2%) | 2 ( 4.3%) | 3 ( 3.3%) | $\chi^2 = 6.49, p = .17, \phi = .27$ |
| High school or GED | 3 ( 6.5%) | 10 (21.7%) | 13 (14.1%) |  |
| Some college/technical | 16 (34.8%) | 16 (34.8%) | 33 (35.9%) |  |
| College (bachelor's) | 14 (30.4%) | 11 (23.9%) | 25 (27.2%) |  |
| Master's or higher | 12 (26.1%) | 6 (13.0%) | 18 (19.6%) |  |
| Employment Category: |  |  |  |  |
| Working (part/full time) | 25 (54.3%) | 16 (34.8%) | 41 (44.6%) | $\chi^2 = 8.54, p = .07, \phi = .30$ |
| Disabled | 5 (10.9%) | 13 (28.3%) | 18 (19.6%) |  |
| Retired | 13 (28.3%) | 9 (19.6%) | 22 (23.9%) |  |
| Other/Declined | 3 ( 6.5%) | 8 (17.4%) | 13 (11.9%) |  |
| Adherence during phase 1 | 97.1% (6.59) | 95.9% (3.09) | 96.5% (5.15) | $t(90) = -1.10, p = .28, d = 0.23$ |
| MVPA during phase 1: |  |  |  |  |
| Week 1 minutes/day | 30.8 (4.17) | 36.6 (4.41) | 33.5 (3.03) | $t(80) = 0.96, p = .34, d = 0.21$ |
| Week 16 minutes/day | 42.1 (23.4) | 49.8 (23.1) | 45.8 (23.5) | $t(80) = 1.50, p = .14, d = 0.34$ |

*Note.* BMI = body mass index; CD4 = cluster of differentiation 4 in white blood cells; GED = general education development degree; MVPA = moderate to vigorous physical activity minutes, intent-to-treat actigraphy data; VACS = Veterans' Aging Cohort Study (comorbidities).

### Procedure

After completing the 16-week in-person, supervised exercise intervention in phase 1 with either high-intensity interval training (HIIT) or continuous moderate exercise (CME), each of which was accompanied by the same resistance exercise training protocol, participants were re-randomized with 1:1 allocation to receive either a tailored-messaging intervention or educational-control messages [17]. The re-randomization sequence was blocked by sex (2 levels), age (3 levels), study site (2 levels), and phase 1 study group (HIIT vs. CME: 2 levels). Messages were digitally delivered to the participant’s device (cellphone or computer) via text, email, or both (participant’s choice). Both conditions first administered a daily survey about barriers to exercise, then delivered daily messages to encourage continued physical activity; however, the tailored-messaging condition had greater use of psychological theory, more message variety, and active tailoring of message content to the participant’s daily survey responses. Participants were encouraged to continue exercising using any method they chose, regardless of their phase 1 group assignment. Participants continued to complete in-person physical function and fatigue assessments every 4 weeks, as they had during phase 1 of the HEALTH study. The study ended as planned in June 2025.

### Interventions

#### Daily surveys

A daily survey was administered to each participant via a link in an SMS text message, email, or both. Surveys asked about 10 possible barriers to exercise commonly reported by PWH. The full text of barrier items from the daily survey is available in Online Resource 1. All participants then immediately received text messages, which differed between experimental groups as described below. Messages in both groups were signed by a HEALTH study team member with a photograph, to increase social motivation for physical activity.

#### Tailored messages

Participants in the tailored-messaging group received MI plus daily text messages that were automatically selected and delivered by an algorithm programmed in the Caspio application development tool (Caspio, Inc.: Sunnyvale, CA). A set of 400 tailored exercise promotion messages was created, and the algorithm selected one of these messages to deliver based on the single most important barrier to exercise that day, according to the individual participant’s daily survey responses. All messages employed evidence-based psychological behavior change strategies selected based on TMT [13], an integrative health behavior model. Sample messages are shown in Online Resource 2. Multiple messages were available for each barrier, with a random component in the algorithm to prevent participants from receiving the same message multiple days in a row. Messages varied in terms of the barrier addressed and the TMT behavior-change strategy used. We also classified messages in terms of whether they included an external URL, provided individualized feedback about the participant’s survey responses, allowed a free-text response, or displayed the participant’s daily MVPA minutes. These features were designed to keep participants engaged by fostering a sense of novelty, which was identified as important in past tailored-messaging research [21]. Surveys and messages were refined based on early feedback from PWH [17].

#### Educational control messages

Participants in the educational-control group received daily messages that were designed to be factual and informative; some also had features such as links to Internet resources on exercise. However, these messages did not use psychological behavior-change principles and were not tailored based on the participant’s survey responses. This condition was designed to control for the “pseudo-tailoring” effect of completing a daily survey and then receiving a message [22], even though there was no actual tailoring in this group. A sample educational message read, “*Keep exercising. Physical activity helps you the most when it’s a long-term habit.*” As with the tailored-messaging intervention, there was a random component to keep participants from receiving exactly the same message multiple days in a row. Nevertheless, there were only 25 total educational-message options due to the lack of personalization, so more message repetition was likely over the course of the study.

#### Coaching using Motivational Interviewing (MI)

Health coaches were trained on MI by the first author, an experienced MI trainer [23]. MI is a well-supported method for promoting health behavior change, including sustaining MVPA [24], using a combination of technical skills (open-ended questions, reflective listening, etc.) and relational empathy [25]. Health coaches used MI during three 5-10 minute conversations with participants, at weeks 16, 20, and 24. MI treatment fidelity was assessed through role-play activities during training, and all health coaches achieved a minimum standard of 80% MI consistency before using this technique with participants. We did not record calls or code MI skills during the intervention itself.

At the University of Colorado site, a health coach delivered MI only to participants in the tailored-messaging group, while participants in the educational-control condition received a check-in call without any use of MI skills. University of Washington health coaches delivered MI to all participants, including those in the educational-control condition as well as those in the tailored-messaging condition (this was a protocol deviation). Therefore, about 50% of participants received tailored messages plus MI, 25% received educational-control messages plus MI, and 25% received educational-control messages without MI. We tested the contribution of MI to the total intervention effect in an exploratory analysis described below.

### Measures

#### Participant Demographic and Clinical Characteristics

PWH provided demographic information before initial randomization. Demographic variables included sex, gender, race, ethnicity, age, education, and employment. Clinical variables were also collected at baseline, including HIV viral load, body mass index, and lowest reported CD4 count (by self-report). A comorbidity metric, the Veterans Aging Cohort Study [VACS] Index, was calculated from baseline laboratory and clinical variables. Throughout the study, adverse events (AEs) and changes in participants’ clinical status were recorded by study staff using a structured form.

#### Primary Outcome

##### Actigraphy-Measured MVPA Minutes per Day

Participants wore an ActiGraph wGT3x-BT monitor (ActiGraph LLC, Fort Walton Beach FL) on their non-dominant hip for four 7-day periods, at study weeks 16, 20, 24, and 28 – i.e., one week per month during phase 2. Participants were asked to wear the ActiGraph during all waking hours; data were considered valid if they included ≥ 4 days with ≥ 10 hours of wear time. Actigraphy data were sampled at a frequency of 30 Hz, with 60-second epochs, and processed using cutoffs from Sasaki et al. [26]: “moderate” activity based on 2690-6166 counts per minute and “vigorous” activity as 6167 counts per minute or more. ActiGraph data are considered valid and reliable for detecting sedentary time, and for differentiating levels of physical activity (light, moderate, vigorous). Participants could not view their own daily results. We summed minutes of moderate and vigorous activity per day to get a daily MVPA total. We then averaged across all available days to calculate weekly MVPA for each participant.

#### Secondary Outcome: Self-Report Minutes of MVPA

We also asked participants to estimate minutes of light, moderate, and vigorous physical activity as part of each daily survey, before the barrier items. Minutes of moderate and vigorous activity were summed to generate an MVPA measure parallel to that obtained from actigraphy. Because self-report data were obtained daily, we averaged across the first 2 weeks for a baseline measurement comparable to the 16-week actigraphy data, then each 4 weeks thereafter to obtain similar timeframes to the 20-, 24-, and 28-week actigraphy data points. Self-report and sensor measures of the same construct often provide complementary information, as they have different sources of error variance [27]. One important source of error in self-reported MVPA was missing data, because we only had MVPA data on days when a survey was completed, and we do not know whether there was a systematic relationship between survey completion and having exercised.

#### Exploratory Measures: Symptoms and Barriers to Exercise

Participants’ daily surveys included yes/no questions about specific barriers; participants who said “yes” were then asked for more detail about that barrier, using established self-report tools. Fatigue was measured using the 4-item Patient-Reported Outcomes Measurement Information System (PROMIS: [28]) fatigue screener, which has been used for daily surveys with PWH, has internal consistency reliability of α = .93 (all alphas in this section were calculated from the current dataset), and correlates with behavioral sleep data as well as biomarkers of inflammation associated with fatigue [29]. Mood was measured using 7 items from the Diary of Ambulatory Behavioral States (DABS: [30]), which has high internal consistency of α = .84 and predictive validity based on prior daily survey research with PWH [29, 31]. Specific task self-efficacy for exercise was measured using a 6-item scale that was also adapted from the DABS [30], with internal consistency reliability of α = .82 and construct validity based on expert consensus and participant review [17]. Pain was measured using the 6-item PROMIS pain screener, which has internal consistency of α = .92 and concurrent validity with functional impairment measures [32]. Motivation was measured using 2 items from the Herzog motivation scale [33], with items selected based on factor loadings in [34]; this scale has internal consistency of α = .84 and predictive validity for behavioral outcomes in PWH [29, 31]. Perceived health was measured using 3 items from the SF-36 [35] with internal consistency of α = .67, including a global health estimate that prospectively predicts mortality in older adults [36]. The daily survey also included six yes/no barrier items adapted from prior research with PWH [31].

### Analysis Plan

All analyses were performed using SPSS 31.0 (Armonk, NY: IBM Corp.). After basic descriptive analysis, we screened for participant characteristics that were used as covariates in phase 1—age, sex, and study site—as potential predictors of MVPA minutes in phase 2. We also tested the potential interaction between phase 2 behavioral intervention and phase 1 exercise condition, in case a specific behavioral approach was more efficacious when paired with either HIIT or CME. For each outcome, we constructed a linear mixed model with time nested within persons, study group as a person-level variable, and the group-by-time interaction effect as the outcome of interest; this is because differences in the pattern of MVPA over time reflect different sustainability of physical activity across groups. All tests were based on fixed effects of predictors, a compound symmetry matrix, person-mean-centering at level 1, and a target □ = .05 for significance. Outcome variables had moderate intra-class correlations of .69 (self-report) to .71 (actigraphy) within persons. To address missing data, we used conservative intent-to-treat (ITT) principles for the main outcome analyses, then also tested complete cases to understand benefits for participants who received the full intervention. In all exploratory analyses, the most conservative ITT-based ActiGraph data were used as the measure of MVPA.

The HEALTH study was powered for the phase 1 test of HIIT versus CME on time to complete a 400-m walk test, which required a sample size of 50 in each condition. We re-randomized participants in phase 2, resulting in half of participants assigned to the tailored-messaging condition and half assigned to the educational-control condition, so there was similar power available for phase 2 as in phase 1, where the target *N* = 100 yielded 88% power to detect effects as small as *d* = 0.63 at □ = .05. As described above, only about a quarter of participants did not receive MI, which led to a lower-powered test of MI’s effects (*d* ≥ 0.81 at power = .80). Our goal for phase 2 was to determine the additional benefit, if any, of tailored messaging for sustained MVPA.

## Results

### Covariate Screening

None of the potential covariates used in phase 1 differed between groups at the phase 2 baseline, as shown in Table 1. Nevertheless, we screened for potential effects of these variables on actigraphy-based MVPA in both the ITT and completer datasets. Only sex predicted MVPA, which was true in both the ITT, *F*(1, 85) = 7.00, *p* = .01, *d* = 0.56, and completers analyses, *F*(1, 83) = 6.41, *p* = .01, *d* = 0.55. Phase 1 group assignment did not predict phase 2 MVPA minutes in either dataset, *F*(1, 84) = 0.35, *p* = .55, *d* = 0.13 for the ITT data. Because age, sex, study site, and phase 1 group assignment were stratification variables for participants’ randomization in phase 1, we nevertheless controlled for these in all models, as well as for main effects when testing interactions; however, a sensitivity analysis showed that models without any covariates led to the same conclusions. We also conducted a sensitivity analysis in which we omitted the 6 participants who exercised independently after phase 1 randomization, in case they differed from those who exercised per the phase 1 protocol; however, all conclusions were unchanged.

### Primary Outcome: Effect on Actigraphy-Measured MVPA

Participants in the tailored-messaging group were more likely to sustain MVPA throughout phase 2, based on a significant group-by-time interaction in the ITT dataset, *F*(1, 255) = 5.76, *p* = .02, *d* = 0.51. Although educational-control participants had slightly higher daily MVPA at the start of phase 2 (*M* = 49.8 minutes, *SD* = 23.1) than those in the tailored-messaging group (*M* = 42.1, *SD* = 23.4), this pattern reversed by 28 weeks as shown in Figure 2 (Panel a), with higher daily MVPA in the tailored-messaging group (*M* = 30.5, *SD* = 36.7) than in the educational-control group (*M* = 26.0, *SD* = 25.8). The completers analysis of actigraphy data similarly showed a significant group-by-time interaction, *F*(1, 274) = 6.57, *p* = .01, *d* = 0.54, again with better maintenance of MVPA minutes over time in the tailored-messaging group than in the educational-control group (Figure 2, Panel b). An exploratory test for the interaction between phase 1 and phase 2 intervention assignment was nonsignificant, *F*(1, 79) = 0.02, *p* = .88, *d* = 0.03 in the ITT dataset, suggesting that tailored messaging was equally effective in promoting sustained MVPA for participants who were initially assigned to either HIIT or CME.

**Fig. 2.**
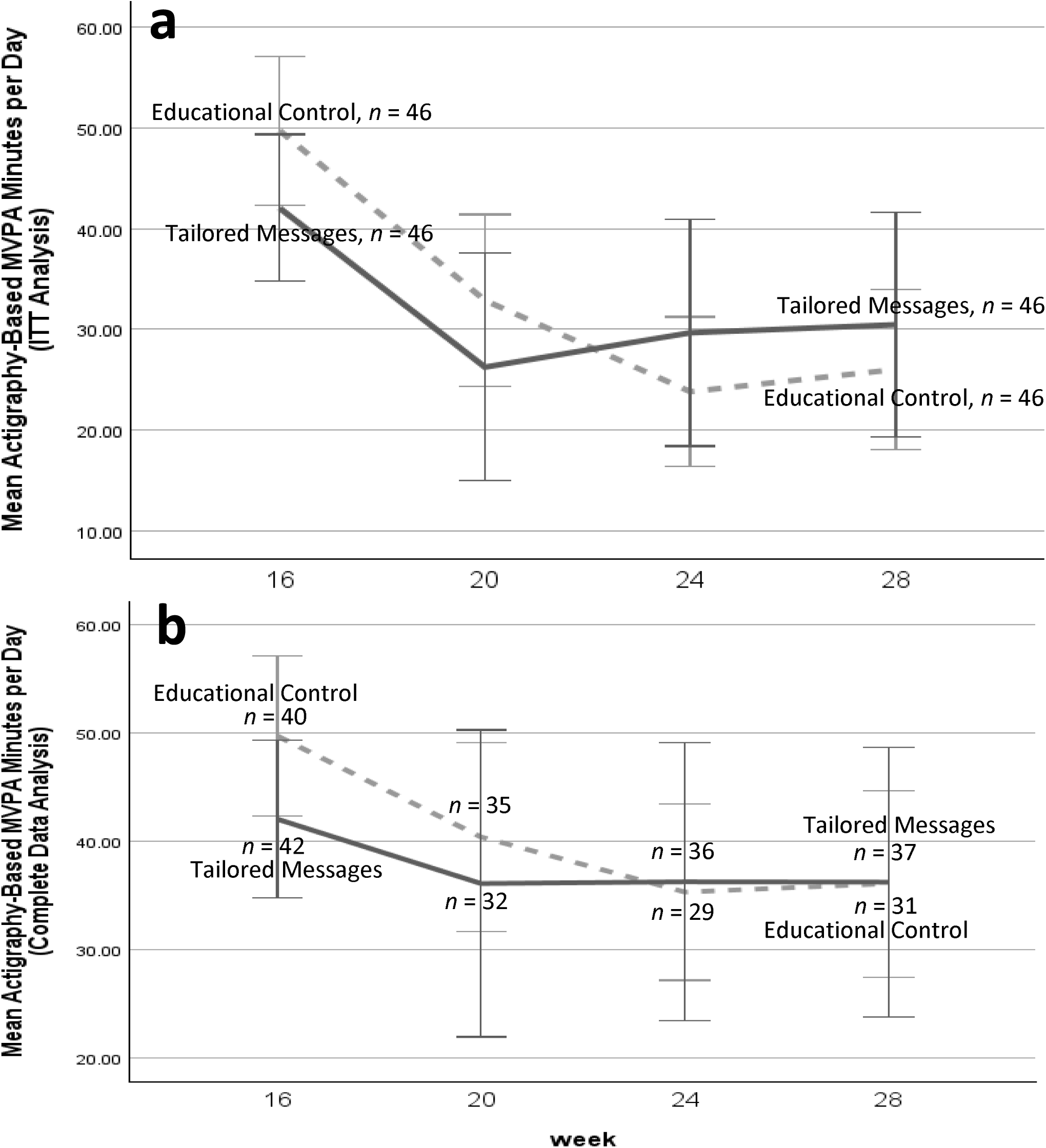
Between-group differences in MVPA minutes per day, based on actigraphy. Panel A shows the intent-to-treat (ITT) analysis, and panel B shows the less-restrictive completers analysis with missing data points omitted. The number of participants with valid actigraphy data, per group at each study week is noted in Figure 2b. The lower number of daily MVPA minutes in the tailored-messaging group (solid line) than in the educational-control group (dotted line) at weeks 16-20 is unexplained, but it appears in both graphs, suggesting that it is not simply a function of missing data. Error bars show ±1.96 standard errors of the mean.

### Secondary Outcome: Effect on Self-Reported MVPA

The analysis of self-reported MVPA minutes confirmed the ActiGraph results, although the overall correlation between the two measures was just *r* = .56. Using an ITT-based dataset in which days with no survey were treated as though there had been no exercise, participants in the tailored-messaging group reported fewer MVPA minutes per day at the start of phase 2 (*M* = 15.9, *SD* = 24.4) than participants in the educational-control group (*M* = 22.7, *SD* = 30.9). This pattern then reversed after 3 months, with better daily MVPA in the tailored-messaging group (*M* = 23.3, *SD* = 42.8) than the educational-control group (*M* = 9.3, *SD* = 13.2). Figure 3 (Panel a) shows the change in ITT-based daily MVPA minutes by month, with a significant group-by-time interaction, *F*(1, 259) = 9.65, *p* = .002, *d* = 0.39. As in the ActiGraph data, there was no interaction between the phase 2 intervention and phase 1 group assignment to HIIT or CME, *F*(1, 82) = 0.73, *p* = .40, *d* = 0.18 based on ITT analysis. Results again might have been biased by attrition, with numbers in Figure 3 (Panel b) showing retention per month in each group. However, the group-by-time interaction remained significant in a completers analysis of self-reported MVPA minutes, *F*(1, 182) *=* 6.76, *p* = .01, *d* = 0.39. Under the plausible assumption that those who continued receiving messages were more adherent to physical activity as well as daily survey completion (in this study, a participant’s average MVPA and the number of surveys they completed were correlated at *r* = +.47), an attrition pattern with more dropout would tend to favor higher MVPA in the control group and a weaker intervention effect. Therefore, our conclusions about self-reported MVPA are robust to challenges based on missing data.

**Fig. 3.**
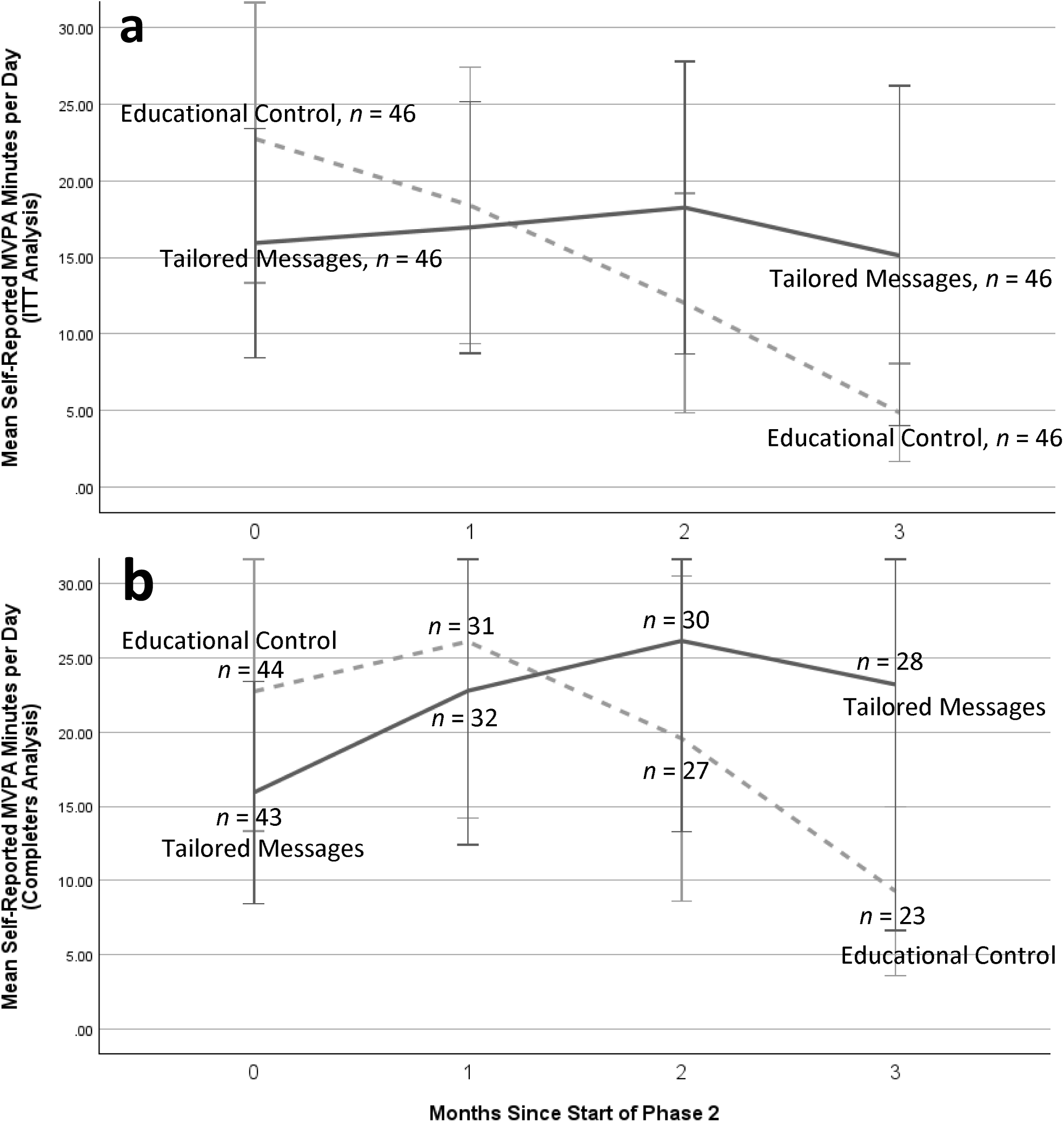
Between-group differences in MVPA minutes per day, based on self-report. MVPA was calculated as the sum of moderate plus vigorous activity minutes reported on daily surveys, and averaged within participants for each month of the phase 2 behavioral intervention. Panel A shows the intent-to-treat (ITT) analysis, where any participant without data was treated as having zero minutes of MVPA. Panel B shows the completers analysis, with averages for those participants who continued to fill out daily surveys; numbers in Figure 2b show how many participants were still receiving messages each month. Solid lines show means for the tailored-messaging group, dotted lines the educational-control group, and error bars ±1.96 SE_Mean_.

### Adverse Events

In phase 2, a total of 10 AEs were reported by 8 participants, all of whom were seen at the Colorado study site. One participant (2 events) was in the tailored-messaging group, with all others in the educational-control group. There were two serious AEs, a COVID-19-related hospitalization and a sinus infection, which were both considered unlikely to be study-related. Two moderate study-related AEs (both in the same participant) involved knee/hip pain and resulted in an emergency room visit. All other moderate AEs were considered unlikely to be study-related, including depression, COVID-19 not requiring hospitalization, flu-like symptoms, flank and leg pain, knee and back pain, and pneumonia. All AEs were resolved with no additional intervention, but in some cases resulted in a missed study visit.

### Intervention Effects on Symptom and Barrier Measures

There were no significant phase 2 intervention effects on the measures of fatigue, mood, pain, motivation for exercise, or any of the following barriers: forgetting, travel, weather, boredom, or “other.” However, there was a significant group-by-time interaction for change in self-efficacy, in which the tailored-messaging group participants reported improved self-efficacy for exercise over time relative to the trend among participants in the educational-control group, *F*(1, 2730) = 4.54, *p* = .03, *d* = 0.08. Similarly, a significant group-by-time interaction for perceived health showed that participants’ scores on SF-36 items stayed at a high level in the tailored-messaging group, while those in the educational-control group decreased, *F*(1, 2765) = 10.4, *p* = .001, *d* = 0.13. Participants in the tailored-messaging group also became less likely to report lack of time as a barrier to exercise, relative to educational-control participants, *F*(2, 2694) = 8.92, *p* < .001, *d* = 0.12 for the group-by-time interaction.

### Exploratory Tests of MI versus Messaging

We conducted an exploratory analysis to determine the contribution of MI to the overall intervention effect documented above. First, we compared sites within the educational-control group, to examine whether MVPA was greater among University of Washington participants (all of whom received MI in addition to daily educational-control messages) than those at the University of Colorado (who did not receive MI in the educational-control group). We excluded tailored-messaging participants from this analysis because all of them had also received MI, so the effects of those two interventions could not be disaggregated. In this test we observed a slight advantage for the educational-control intervention at the University of Washington, *M* = 43.8 MVPA per day (*SD* = 2.59, *N* = 23) versus the University of Colorado, *M* = 36.3 (*SD* = 21.0, *N* = 23), suggesting that MI was a beneficial addition to the educational-control messages, even though this test did not reach conventional levels for statistical significance, *F* (1, 37) = 1.48, *p* = .23, *d* = 0.37. We then looked at University of Washington participants alone, all of whom received MI, with *N* = 24 in the tailored-messaging group and *N* = 23 in the educational-control group. We found an even stronger effect of tailored messaging (with MI) compared to the educational-control messages (with MI), *F*(1, 130) = 15.1, *p* < .001, *d* = 1.16 for the group-by-time interaction effect. The second of these exploratory tests suggests that tailored messaging had a positive effect independent of MI. But because of the site-by-condition confounding, this exploratory analysis could not conclusively determine whether MI or tailored messages (or both) was the active ingredient in the MI plus tailored-messaging group.

### Other Exploratory Analyses

Given the variety of strategies to explain sustained MVPA that were reviewed in the Introduction, we also conducted exploratory analyses to determine whether specific types of intervention messages were more effective than others (Table 2). Participants reported significantly higher MVPA minutes on days when they were able to enter a free-text response to that day’s message, when the message emphasized either mindfulness or social connection, and when the participant reported any barrier other than lack of time for exercise. These effects were particularly evident in the tailored-messaging group, with minimal differences among educational-control participants. Other broad intervention characteristics such as whether the message contained an external URL (e.g., to a video or interactive resource), displayed the participant’s current active minutes, or provided other personalized feedback from the daily survey, were not significantly associated with daily MVPA. It is still possible that some of these features were more effective for specific participants or at specific times, a matching hypothesis that we plan to explore in future research.

**Table 2.**
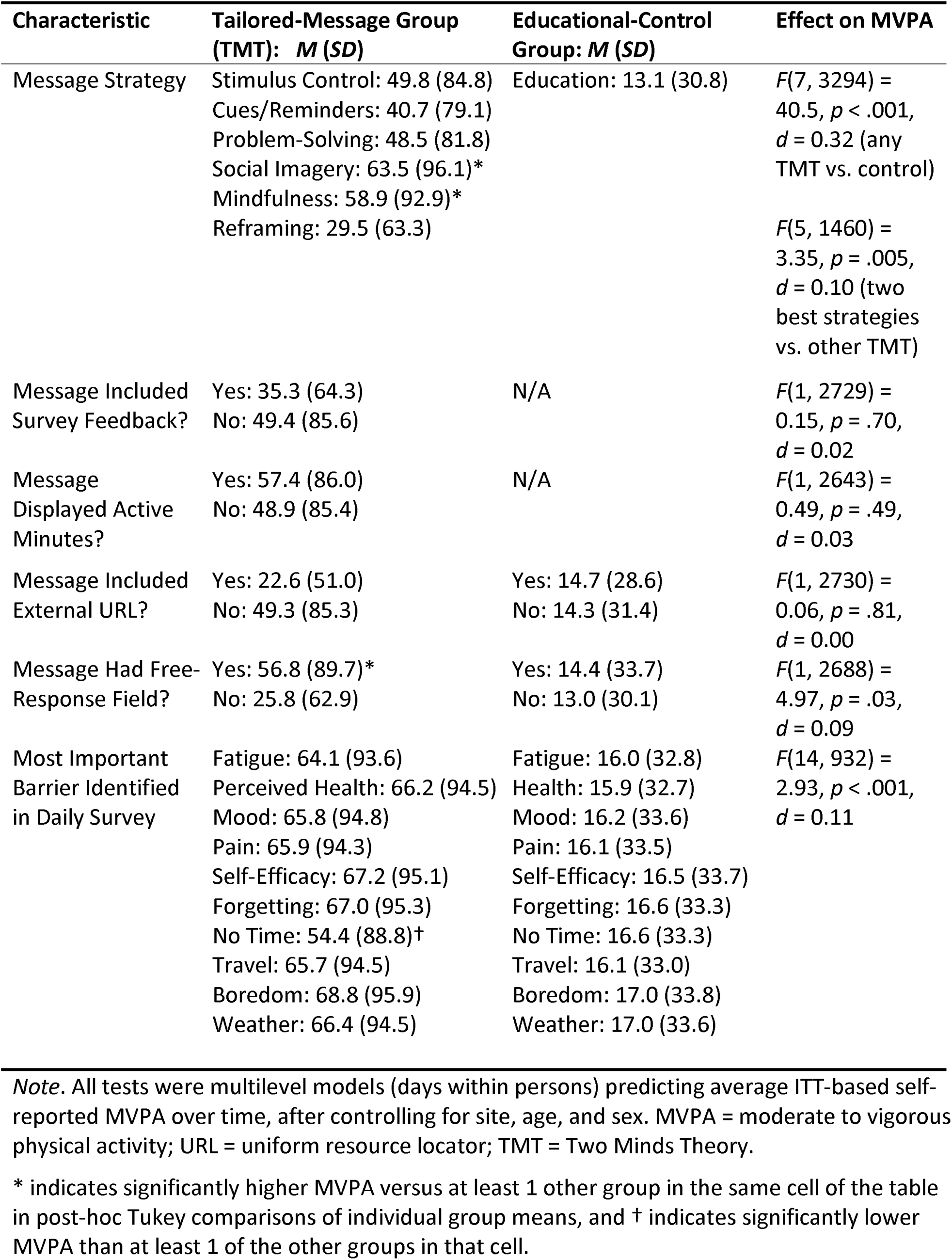
Effect of Aggregate Message Characteristics on Daily MVPA Minutes.

## Discussion

This randomized trial showed that psychologically tailored messages based on TMT were efficacious for sustaining MVPA among older PWH, in comparison to non-tailored educational messages delivered by the same automated system, on the same schedule, and in response to the same daily survey. Results were similar whether daily MVPA minutes were measured by actigraphy or self-report; even though both measures were affected by attrition to some degree, convergent results suggest a positive effect. Furthermore, using ITT principles with the very conservative assumption that people without data were not exercising at all still showed a positive effect. This study did not include a no-treatment comparison, so we do not know if educational-control participants’ results mirror what would have happened in the absence of any intervention.

The effect of tailored messages was potentially separate from the effect of MI delivered by an exercise coach, which was also part of this multi-component intervention, based on evidence from the site that delivered MI to both groups. At that site, the MI-plus-tailored-messaging intervention was still superior to the MI-plus-educational-messaging condition. By comparing across study sites, we estimated the effect of MI alone to be *d* = 0.37, while the MI-plus-tailored-messaging effect was *d* = 0.51, suggesting that about 27% of the combined effect was due to tailored messaging. MI is an intervention with mixed evidence for physical activity promotion and maintenance, with some studies showing positive results [12, 24, 37] but others showing limited benefits [7, 38]. The effect of MI alone cannot be confidently determined from this study based on design considerations, and it is possible that the proportion of the total intervention effect due to MI was either higher or lower: We note that the effect size of MI in this study was lower than that in the meta-analysis of MI for exercise by Zhu et al. [37], *d* = 0.45, but that MI has not consistently shown benefits for exercise [7], and that our estimated effect of *d* = 0.37 was also higher than the average effect of MI across health behaviors reported by Lundahl et al. [24], *g* = 0.28. Thus, although both MI and tailored messaging seem to have had positive effects, further study is needed to determine their relative contributions.

In exploratory tests, participants in the tailored-messaging group also reported more self-efficacy for exercise over time, better perceived health, and less of a perception that they did not have time to exercise, relative to the trend among educational-control participants. The self-efficacy change is consistent with one of the psychological mechanisms linked to exercise maintenance in prior research [9]. However, the current analysis did not test mediation, and the results should be interpreted as showing that a TMT-based intervention improved self-efficacy, rather than showing that self-efficacy changes explained the effects of TMT-based messaging. The perceived health findings could be related to PWH maintaining a higher level of fitness as they continued to exercise. These exploratory results suggest that an MVPA sustainability intervention can potentially lead to benefits beyond cardiovascular fitness.

Finally, with regard to behavioral theory, some message characteristics were associated with higher daily MVPA than others. For example, messages focused on the barrier of time were associated with a slightly lower level of MVPA than messages targeted to other barriers. Messages that offered the opportunity for a free-text response were associated with higher same-day MVPA minutes. However, these message characteristics did not produce any differential benefit in the educational-control group. All individual TMT-based messaging strategies were effective compared to educational-control messages, but among the TMT messages there was some advantage for those based on mindfulness or social imagery. This finding might help to further refine the TMT algorithm for delivering specific messages to specific patients, and further research on person-message matching is planned. TMT is an integrative model that incorporates effective behavior-change strategies from many existing theoretical traditions [13], so it is possible that others could see our results as supporting other behavior-change models – e.g., Bandura’s Social Cognitive Model [9] would be consistent with the social-imagery messaging strategy, and with the observed increase in self-efficacy. Ultimately, we believe the strength of TMT is in its focus on everyday situational factors, such as barriers to exercise, and how these factors change within persons over time. TMT’s time-based within-person focus makes it a particularly applicable theory for daily tailored-messaging interventions. Further research is needed to develop more fine-grained matching hypotheses, and to directly compare TMT-based interventions to those based on other theoretical models.

Although our educational-control group included some features designed to create a perception that those messages were also custom-selected (to avoid a “pseudotailoring” effect), it is still possible that people’s appreciation for the tailoring itself contributed to the intervention effect [22]. A second possible explanation is that tailored messages offered more variety to maintain participants’ attention; the educational-control group had fewer total messages available, so statistically each one was likely to come up more often, which might have led to enhanced message features being seen as more novel in the tailored-messaging group [21]. The lower rate of attrition in the tailored messaging group is consistent with a novelty-based interpretation. Nevertheless, all tailored message types were effective, and any differences among message types were small (*d* = 0.10) compared to the overall effect of the tailored-messaging intervention (*d* = 0.51). Future research could examine contextual or person-level differences that might moderate the efficacy of specific message characteristics.

### Limitations and Directions for Future Research

Positive intervention effects were seen both in completers and ITT analyses, suggesting that our findings cannot be explained by attrition alone. Similarly, the results were consistent across both self-report and actigraphy measures of MVPA, which suggests that our findings are not the result of self-report bias or selective recall. Despite these strengths of the study, attrition and self-report bias each remain potential weaknesses, which might have affected our results in unknown ways. Because both the actigraphy and self-report measures had missing data, this study had no single “gold standard” measure of MVPA. Despite that weakness, the consistency of findings across various measures still suggests a positive intervention effect.

Our exploratory subgroup analysis focused on MI suggests that tailored messages could have an independent effect. However, this test was perfectly confounded with intervention site (one of the sites delivered MI to educational-control-group participants and the other did not), and participants did differ in important ways by site, so it is possible that other site-level differences affected the results. Although we interpret our findings as an additive effect of MI plus tailored messages, it is also possible that the MI and tailored-messaging components interacted in some way, or that one or the other of the intervention components was not needed. Our estimate for the contribution of MI is slightly higher than meta-analytic findings about the average effect of MI for exercise promotion, which might mean that the tailored messaging component was more important than our current estimate, but it is also true that MI was always part of the intervention and that we do not know whether tailored messages alone would have any benefit. Future research with a group that receives *only* tailored messages is needed to determine whether tailored messaging can help to sustain MVPA on its own.

Findings about the intervention effect on other outcomes were exploratory, and the perceived health scale in particular had sub-optimal internal consistency reliability (*α* = .67). Between-group differences on variables other than MVPA should be viewed with caution and are in need of replication. Additionally, there are clear generalizability limitations for our findings: Although our participants were representative of PWH in the Western United States, proportionally more PWH in this section of the country are White men who have sex with men, compared to the U.S. HIV epidemic which has greater representation of women and minority group members. Additional studies involving more diverse PWH in other parts of the country are needed to determine whether our results are limited to a specific sub-population of PWH.

Finally, we again note that all participants completed a 16-week structured exercise intervention before starting this behavioral intervention trial, and that messages in this study were designed to promote sustained MVPA among people who were already exercising. This study therefore does not provide any evidence on the question of whether tailored messages would help people to initiate a new exercise program, or to establish a regular exercise habit during the first 3 months. In Prochaska et al.’s [6] framework, the first 3 months of a new behavior are considered the “action phase,” separate from the later “maintenance phase” when exercise has already become a habit. Different interventions may be required for these two distinct steps in the process of health behavior change.

### Conclusion

In this randomized trial, following 16 weeks of supervised exercise, PWH had more sustained MVPA when they received an intervention consisting of tailored messages plus MI. This was true even though the comparison group also received an intervention that included daily self-assessment surveys, educational messages, and in some cases MI. Because sustaining MVPA over the long term is critically important for maintaining its health benefits, this type of low-cost intervention should be considered for PWH who successfully initiate exercise. Future research could investigate the applicability of this intervention to people who are starting a new exercise habit, to people with other chronic conditions, or to older adults in general.

## Statements and Declarations

### Compliance with Ethical Standards

*Ethics Approval:* This study was approved by the Colorado Multiple Institutional Review Board, protocol #19-1985. *Pre-Registration:* ClinicalTrials.gov study #NCT04550676. *Informed Consent:* All participants provided written informed consent.

### Data Availability

De-identified participant data with a data dictionary will be available upon reasonable request to the corresponding author (PFC), upon institutional review board approval, with additional agreement, and dependent on approval of a data analysis protocol.

### CRediT Statement

Administration: ARW, KME; Conceptualization: PFC, ARW, SM, CMJ, KME; Data curation: PFC; Formal analysis: PFC, MPW, SM; Funding: ARW, KME; Investigation: VHFO, VK, GLK; Methodology: PFC, ARW, MPW, SM, SMaW, CMJ, KME; Supervision: ARW, KME; Visualization: PFC; Validation, review and editing: all authors; Writing – original draft: PFC.

