## Supplemental Table 1 for "Efficacy of Tailored Messages for 28-Week Exercise Sustainability in People with HIV"

**Supplementary Table 1.** Description of Daily Survey Items

| *Barrier* | *Screening Item (yes/no)* | *Detail Items* |
| --- | --- | --- |
| Fatigue | Since the last time you took this survey, have you been too tired to exercise? | Modified PROMIS fatigue screener (1-5 scale),^[1]^ with the wording “since the last time you took this survey” instead of “over the last 7 days.” |
| Perceived Health | Since the last time you took this survey, has your health gotten in the way of exercising? | One item from the SF-36 (5-point scale),^[2]^ plus two new items about the effects of health on exercise (felt too sick, skipped exercise because I felt fine) created by our team (4-point scale). |
| Self-Efficacy | Since the last time you took this survey, have you felt like you weren't able to exercise? | Two items from DABS control measure (4-point scale);^[3]^ plus four from the SCI Self-Efficacy for Exercise^[4]^ measure of exercise-specific self-efficacy, using the 4-point DABS response scale. |
| Mood | Since the last time you took this survey, has your mood gotten in the way of exercising? | Seven items: four from the DABS mood scale^3^ and three others created by our team in the same format (4-point scale). |
| Pain | Since the last time you took this survey, has pain gotten in the way of exercising? | Six-item PROMIS pain screener (1-5 scale),^[5]^ with the wording “since the last time you took this survey” instead of “over the last 7 days.” |
| Forgetting | Since the last time you took this survey, have you forgotten to exercise? | N/A |
| Competing Priorities | Since the last time you took this survey, did you skip exercise because you didn’t have the time? | N/A |
| Boredom | Did you skip exercise because it was too boring? | N/A |
| Travel | Did you miss exercise because you were away from home? | N/A |
| Weather | Did you skip exercise because the weather wasn’t good for it? | What type of weather was the problem?  Rain or Snow Too Hot Too Cold  Too Windy Poor Air Quality  Technical Difficulties (e.g., broken treadmill, gym closed, wifi down) Other |
| Other | Are there other barriers to exercise that you would like to let us know about? | What was the other barrier to exercising?  [Free-text response] |

*Note*. The survey used branching logic to present supplemental questions in the “detail items” category only if the screening item for that barrier was marked “yes.”

DABS = diary of ambulatory behavioral states; PROMIS = patient-reported outcomes monitoring system

1. PROMIS 4-item fatigue screener: Cella D, Choi, S. W., Condon, D. M., Schalet, B., Hays, R. D., Rothrock, N. E., ... & Reeve, B. B. PROMIS® adult health profiles: efficient short-form measures of seven health domains. *Value in Health*. 2019;22(5):537-544. Retrieved from <https://www.healthmeasures.net>. Adapted wording from Makic, M. B., Gilbert, D., Jankowski, C., Reeder, B., Al-Salmi, N., Starr, W., & Cook, P. F. (2020). Sensor and survey measures associated with daily fatigue in HIV: findings from a mixed-method study. *Journal of the Association of Nurses in AIDS Care*, *31*(1), 12-24. https://doi.org/10.1097/JNC.0000000000000152
2. SF-36 item 1 measuring subjective health retrieved from <https://www.rand.org/health-care/surveys_tools/mos/36-item-short-form/survey-instrument.html>
3. Kamarck, T.W., Shiffman, S., Smithline, L., Goodie, J., Thompson, H., Ituarte, P.H., Jong, J., Pro, V., Paty, J., Kassel, J., Gnys, M. & Perz, W. (1998). The Diary of Ambulatory Behavioral States: A New Approach to the Assessment of Ambulatory Cardiovascular Activity (pp. 163-193). In D. Krantz & A. Baum (Eds.): *Perspectives in Behavioral Medicine: Technology and Methodology in Behavioral Medicine*. Hillsdale, N.J.: Lawrence Erlbaum.
4. Kroll, T., Kehn, M., Ho, P. S., & Groah, S. (2007). The SCI exercise self-efficacy scale (ESES): development and psychometric properties. *International Journal of Behavioral Nutrition and Physical Activity*, *4*(1)*,* 34. <https://doi.org/10.1186/1479-5868-4-34>
5. PROMIS 6-item pain interference short form: Kim J, Chung H, Amtmann D, Revicki DA, Cook KF (2013). Measurement invariance of the PROMIS pain interference item bank across community and clinical samples. *Quality of Life Research, 22*(3), 501-7. Retrieved from <https://www.healthmeasures.net>. Adapted wording from Makic, M. B., Gilbert, D., Jankowski, C., Reeder, B., Al-Salmi, N., Starr, W., & Cook, P. F. (2020). Sensor and survey measures associated with daily fatigue in HIV: findings from a mixed-method study. *Journal of the Association of Nurses in AIDS Care*, *31*(1), 12-24. <https://doi.org/10.1097/JNC.0000000000000152>
