## Supplemental Table 2 for "Efficacy of Tailored Messages for 28-Week Exercise Sustainability in People with HIV"

**Supplementary Table 2.** Barriers, Behavior Change Principles, and Sample Results of the Tailored Messaging System

| *Barrier to Exercise* | *Principle of Behavior Change^1^* | | *Sample System-Generated Message* |
| --- | --- | --- | --- |
| Negative Mood | Intuitive 1 (behavioral economics): leverage biases and heuristics  Narrative 1 (social imagery): imagine emotional consequences of actions | “People’s mood often improves if they spend time outdoors. What simple outdoor activity would get you moving?”*  “What might you tell a good friend who wanted to get more exercise? What advice would you give to motivate them?”* | |
| Fatigue | Intuitive 2 (behaviorism): train behavior using cues and prompts  Narrative 2 (mindfulness): attend to internal states for motivation | “When do you usually exercise? If you have trouble falling asleep, make it a habit to exercise earlier in the day.”  “After you exercise, notice whether you feel more tired or less. Does exercise wear you out? Give you energy? Both?”* | |
| Low Self-Efficacy | Intuitive 3 (problem-solving): seek unfiltered creative ideas, then focus  Narrative 3 (cognitive therapy): reframe current experiences | “Make a list of 5 kinds of exercise you might want to try. Pick one to try. Tomorrow try another. See which you like better.”  “Exercise can give you a sense of control, when the rest of your life feels out of control.” | |
| Poor Health | Intuitive 2 (behaviorism): train behavior using cues and prompts  Narrative 3 (cognitive therapy): reframe current experiences | “Not feeling up to a high-intensity workout? Try a 10-minute yoga video instead: [link to external video here]”†  “Football coach Vince Lombardi said, ‘it’s not whether you get knocked down, it’s whether you get up.’ What can you do to get back up and be active today?” | |
| Time/Priorities | Intuitive 3 (problem-solving): seek unfiltered creative ideas, then focus  Narrative 2 (mindfulness): attend to internal states for motivation | “Think about how you spent time yesterday. Were some activities less essential? Could you use that time to exercise?”  “Do you get more done on days when you exercise, or less? Are there ways exercising helps you get other things done?”* | |
| Pain | Intuitive 3 (problem-solving): seek unfiltered creative ideas, then focus  Narrative 1 (social imagery): imagine emotional consequences of actions | “What has helped you cope with pain in the past? How can you use any of those strategies to get moving again now?”*  “If you know someone else who struggles with pain, ask how they manage it. Do they have strategies you’d like to try?” | |
| Forgetting | Intuitive 1 (behavioral economics): leverage biases and heuristics  Narrative 2 (mindfulness): attend to internal states for motivation | “Try setting out your exercise clothes at night, so that in the morning you’re ready to jump in them and go.”  “Notice what’s different on days when you remember to exercise versus when you forget. What can you learn from the experience of forgetting? How will that help you next time?”* | |
| Travel | Intuitive 2 (behaviorism): train behavior using cues and prompts  Narrative 2 (mindfulness): attend to internal states for motivation | “Try to get outside for some fresh air or a break. Walk to lunch or dinner instead of staying close to your hotel.”  “Travel can be stressful! Take a look at your mood score for today: [score]. Exercise is a great form of stress relief.”‡ | |
| Boredom | Intuitive 3 (problem-solving): seek unfiltered creative ideas, then focus  Narrative 1 (social imagery): imagine emotional consequences of actions | “What strategies could you try to stay motivated for exercise? Write down a list.”*  “Exercise can be more interesting when there’s an element of competition. Try competing with a friend to see who can get more steps next week!” | |
| Environment  (6 subtypes: hot, cold, wet/windy, air quality, resource gap,^2^ other) | Intuitive 1 (behavioral economics): leverage biases and heuristics  Narrative 3 (cognitive therapy): reframe current experiences | “If you can’t exercise outdoors, try an indoor cardo workout instead: [link to external video here]”†  “Did you know that your body burns more calories when it’s cold? This is a chance to be really efficient in your exercise!” | |

^1^ Examples only. Note that all barriers had possible messages connected to all 6 theory-based strategies, resulting in a 10x6 matrix of possible barriers crossed with possible strategies, and usually multiple messages within each cell of that matrix. There are 3 possible Intuitive-Mind strategies and 3 possible Narrative-Mind strategies.

^2^ The “resource gap” category of environmental barriers included events like “gym closed” or “fitness tracker broken.”

* Shows examples of tailored messages that allowed the participant to enter a free-text response.

† Shows examples of tailored messages that included a web link to a video or other external resource.

‡ Shows example of a tailored message that included feedback from one of the participant’s survey responses that day.
